# Declining Adherence to Dietary Approaches to Stop Hypertension (DASH) Diet Among US Adults with Hypertension: NHANES 2003–2018

**DOI:** 10.64898/2026.09.28.26364241

**Authors:** Vijaya Parameswaran, Anjali Phalke, Christopher Gardner, David J. Maron

## Abstract

**Background:** Data from National Health and Nutrition Examination Survey (NHANES) cycles 1988–1994 and 1999–2004 show a declining trend in the adherence to Dietary Approaches to Stop Hypertension (DASH) among adults. Since then, DASH evidence, dietary guidelines, and public-facing dietary resources have advanced, but whether national DASH adherence has improved is uncertain.

**Methods:** We extended the Mellen et al. NHANES analysis among 14,163 adults with self-reported hypertension (N=6,914, 2003–2010, N=7,249, 2011–2018). DASH score included 9 nutrient components: total fat, saturated fat, protein, cholesterol, fiber, calcium, magnesium, potassium, and sodium, with adherence defined as a score ≥4.5. Survey-weighted multivariable models compared DASH score, adherence, and nutrient-threshold attainment across periods.

**Results:** In 2011–2018, U.S. adults with self-reported hypertension represented 76.2 million individuals (33.8%). Multivariable-adjusted DASH adherence declined from 22.1% in 2003-2010 to 18.6% in 2011-2018 (p=0.003), and mean DASH score declined from 2.98 to 2.85 (p=0.007). Adjusted odds of adherence were lower in 2011–2018 compared with 2003–2010 (OR, 0.78; 95% CI, 0.67–0.91; p=0.002). Adherence was lower among non-Hispanic Black adults (OR, 0.70; 95% CI, 0.59–0.84), adults with BMI ≥30 kg/m² (OR, 0.61; 95% CI, 0.51– 0.73), and adults with prior stroke (OR, 0.71; 95% CI, 0.56–0.89). Total fat and sodium threshold attainment declined from 20.9% to 18.7% and 13.8% to 11.8%, respectively.

**Conclusions:** DASH adherence among U.S. adults with self-reported hypertension declined from 2003-2011 to 2011-2018. Implementation strategies should identify barriers to adoption and address the full DASH dietary pattern.

## INTRODUCTION

Hypertension, defined as systolic blood pressure (SBP) ≥130 mmHg or diastolic blood pressure (DBP) ≥80 mmHg, affects nearly half of U.S. adults and is a leading modifiable risk factor for cardiovascular disease, yet only one-quarter achieve adequate blood pressure control[1–4]. The Dietary Approaches to Stop Hypertension (DASH) diet rich in fruits, vegetables, and low-fat dairy and reduced in saturated fat and cholesterol is among the most effective non-pharmacological approaches to lowering blood pressure (BP)[5–9]. Meta-analyses show that DASH lowers SBP and DBP by an average of 6.74 mm Hg and 3.54 mm Hg, respectively, with additional benefits for body weight, lipid profiles, and long-term risk of coronary heart disease and stroke [7]. Despite this evidence, using data from the National Health and Nutrition Examination Survey (NHANES) III, 1988–1994, and NHANES 1999–2004, Mellen et al. reported that DASH adherence among adults with hypertension declined from 26.7% to 19.4%, largely reflecting fewer adults meeting targets for total fat, fiber, and magnesium, suggesting that dissemination of dietary evidence had not translated into meaningful population-level dietary change [12]. Since then, U.S. dietary guidelines have increasingly emphasized heart healthy overall dietary patterns and national sodium reduction initiatives have been introduced[13–19]. Whether these shifts have influenced population-level DASH adherence among the subset of Americans with elevated blood pressure who would benefit the most remains unknown. We therefore extend the Mellen et al. analysis to characterize trends in DASH adherence among U.S. adults with self-reported hypertension using NHANES data from 2003 through 2018.

## METHODS

### Survey Overview and Analytic Sample

This study used data from the National Health and Nutrition Examination Survey (NHANES), a series of cross-sectional surveys representing the civilian, non-institutionalized US population through a complex, multistage probability design. The primary analysis used continuous NHANES cycles from 2003–2004 through 2017–2018. The analytic sample included adults aged ≥20 years who completed the household interview, attended the mobile examination center, completed a single 24-hour dietary recall, and had nonmissing day-1 dietary sample weights. Pregnant participants were excluded. Participants were also required to have nonmissing data needed to define hypertension status, including self-reported hypertension status and measured blood pressure among those without self-reported hypertension. The descriptive hypertension-status analysis included 38,024 adults and categorized participants as normotensive, prehypertensive, having undiagnosed hypertension, or having self-reported hypertension. The primary analytic cohort comprised 14,163 adults with self-reported hypertension, defined as an affirmative response to whether a physician or health professional had ever told the participant they had hypertension or high blood pressure.

### Demographics and Physical Examination

Demographic variables included age, sex, race/ethnicity, educational attainment, poverty-income ratio (PIR), and health insurance status. Age was categorized as 20–39, 40–59, and ≥60 years. Race/ethnicity was harmonized using RIDRETH1, the cross-cycle NHANES recode available across 2003–2018. Education was categorized as less than high school, high school or GED, and more than high school. PIR was categorized as <1, 1–2.9, 3–4.9, and ≥5.

Blood pressure was measured at the mobile examination center. Participants were grouped into four mutually exclusive categories: self-reported hypertension (prior physician diagnosis); undiagnosed hypertension (mean systolic blood pressure [SBP] ≥140 mm Hg or mean diastolic blood pressure [DBP] ≥90 mm Hg among those without self-reported hypertension); prehypertension (mean SBP 120–139 mm Hg or mean DBP 80–89 mm Hg); and normotension (mean SBP <120 mm Hg and mean DBP <80 mm Hg). When multiple blood pressure readings were available, the first was excluded and the remainder averaged; when only one reading was available, that value was used. Body mass index (BMI) was calculated from measured height and weight and categorized as <25.0, 25.0–29.9, and ≥30.0 kg/m². Medical history variables were based on self-reported physician or health professional diagnosis, consistent with the approach used by Mellen et al., and included diabetes, high cholesterol, coronary heart disease, stroke, and congestive heart failure.

### Dietary Assessment and DASH Score

Dietary intake was assessed using a single24-hour dietary recall administered at the mobile examination center. A nine-component DASH nutrient score was constructed from foods and beverages based on the framework reported by Mellen et al., encompassing total fat, saturated fat, protein, cholesterol, fiber, calcium, magnesium, potassium, and sodium. Macronutrients were expressed as percentage of total energy intake; remaining nutrients were indexed per 1000 kcal. Each component was scored 1.0 point for meeting the full DASH target, 0.5 points for meeting an intermediate target, or 0 otherwise, and participants with a total score ≥4.5 were classified as DASH adherent [12].

### Scoring Approach

To ensure comparability with the original publication, we validated the scoring approach in NHANES 1999–2004, reported by Mellen et al. We found that although our results closely matched the published values for nutrient estimates, applying the reported 0.5-point intermediate scoring produced lower DASH scores than those published. Therefore, a calibrated intermediate score of 0.75 was used in the primary analysis as it reproduced the original estimates more closely and a sensitivity analysis using the 0.5-point intermediate score was conducted.

### Period Definitions

The primary analysis compared DASH adherence between 2003–2010 and 2011–2018. Descriptive hypertension-status tables and subgroup analyses used the pooled 2003–2018 dataset. A secondary historical-context analysis used four non-overlapping periods: NHANES III 1988–1994, NHANES 1999–2004, 2005–2010, and 2011–2018. This approach avoided double-counting the 2003–2004 cycle across both the historical and primary analysis periods.

### Statistical Analysis

All analyses accounted for the NHANES complex survey design using strata, primary sampling units, and single 24-hourdietary sample weights. Dietary weights were divided by 8 for pooled 2003–2018 analyses, by 4 within each 8-year period for the primary comparison, by 3 for the 2005–2010 historical period, and by 4 for 2011–2018. Descriptive characteristics were estimated as survey-weighted means or percentages with standard errors. DASH score was analyzed using survey-weighted linear regression; DASH adherence and individual nutrient-target attainment were analyzed using survey-weighted logistic regression and reported as adjusted predicted percentages with standard errors. Models were first adjusted for age category and total energy intake, then additionally adjusted for sex, race/ethnicity, education, PIR, and diabetes. Multivariable logistic regression was used to estimate odds ratios for DASH adherence. Missing covariate data were handled using complete-case analysis. Age- and energy-adjusted models included all 14,163 adults with self-reported hypertension and nonmissing dietary data. Multivariable models for adjusted DASH score, DASH adherence, and nutrient-target attainment included 12,966 participants with complete data for survey period, age category, total energy intake, sex, race/ethnicity, education, poverty-income ratio, and diabetes mellitus. The expanded multivariable logistic regression model estimating odds ratios for DASH adherence additionally required complete data for body mass index, high cholesterol, coronary heart disease, stroke, and congestive heart failure and included 11,747 participants. Analytic sample sizes and complete-case exclusions by model are summarized in Supplemental Table S3. Two-sided p values <.05 were considered statistically significant. Analyses were conducted in R version 4.4.0 using the survey package.

**Table 1.**
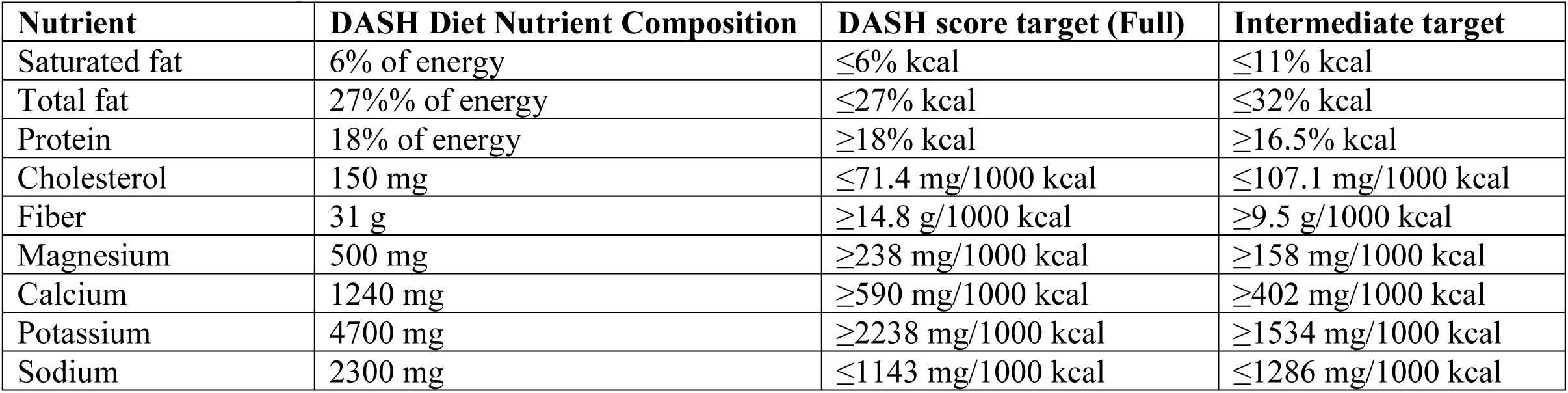
Nutrient targets for DASH score. Abbreviation: DASH. Dietary Approaches to Stop Hypertension trial. Dash composition based on a 2100-kcal diet. Full targets contributed 1.0 point in the primary validation-calibrated score. Intermediate targets derived from Mellen et. al (2008), contributed 0.75 points in the primary score and 0.5 points in the sensitivity score.

## RESULTS

### Hypertension Status and Study Population

Across NHANES 2003–2018, the adult sample was distributed across hypertension-status categories as follows: 37.7% normotensive, 23.2% prehypertensive, 6.4% with undiagnosed hypertension, and 32.8% with self-reported hypertension (Table 2). The analytic sample represented approximately 64.5 million U.S. civilian, noninstitutionalized adults with self-reported hypertension in 2003–2010, corresponding to 31.7% of U.S. adults, and 76.2 million adults in 2011–2018, corresponding to 33.8% of U.S. adults. Sociodemographic differences were observed across hypertension-status groups, and dietary differences were most apparent for cholesterol and sodium density, both of which were higher among adults with self-reported hypertension than among normotensive adults (cholesterol: 143 vs. 134 mg/1000 kcal; sodium: 1725 vs. 1677 mg/1000 kcal). In adjusted models, DASH score differed significantly across hypertension-status groups (Supplemental Table S1), with the lowest score observed among adults with self-reported hypertension.

**Table 2.** Demographic, clinical, and dietary characteristics by hypertension status, NHANES 2003-2018.

| **Characteristic** | **Normotensive**<br>**(N=12911)** | **Prehypertension**<br>**(N=8288)** | **Undiagnosed**<br>**hypertension**<br>**(N=2662)** | **Self-reported**<br>**hypertension (N=14163)** |
| --- | --- | --- | --- | --- |
| Population % | 37.7 (0.5) | 23.2 (0.4) | 6.4 (0.2) | 32.8 (0.5) |
| Women % | 58.7 (0.5) | 40.0 (0.8) | 43.6 (1.4) | 51.9 (0.6) |
| Age in years:% (SE) |  |  |  |  |
| 20-39 | 56.8 (0.9) | 38.5 (0.9) | 18.0 (1.1) | 13.4 (0.5) |
| 40-59 | 33.3 (0.7) | 42.1 (0.9) | 40.9 (1.4) | 38.9 (0.6) |
| >=60 | 9.9 (0.5) | 19.4 (0.7) | 41.1 (1.4) | 47.7 (0.8) |
| Race/ethnicity: % (SE) |  |  |  |  |
| Non-Hispanic White | 66.7 (1.3) | 67.4 (1.3) | 68.9 (1.8) | 70.2 (1.3) |
| Non-Hispanic Black | 9.2 (0.5) | 10.8 (0.7) | 12.2 (1.0) | 13.9 (0.9) |
| Mexican American | 10.3 (0.7) | 9.2 (0.7) | 8.0 (0.9) | 5.4 (0.5) |
| Other Hispanic | 6.0 (0.4) | 5.1 (0.5) | 4.2 (0.5) | 4.1 (0.4) |
| Other Race/Multi-Racial | 7.8 (0.5) | 7.5 (0.5) | 6.7 (0.7) | 6.4 (0.4) |
| Education: % (SE) |  |  |  |  |
| <High school | 13.7 (0.6) | 14.9 (0.6) | 18.5 (1.1) | 18.7 (0.6) |
| High school/GED | 21.7 (0.7) | 24.4 (0.8) | 26.5 (1.2) | 25.7 (0.6) |
| >High school | 64.6 (1.0) | 60.7 (1.1) | 55.1 (1.5) | 55.6 (0.9) |
| Poverty income ratio: % (SE) |  |  |  |  |
| <1 | 15.8 (0.6) | 12.6 (0.6) | 12.2 (0.9) | 13.6 (0.5) |
| 1-2.9 | 34.1 (0.8) | 34.3 (1.0) | 38.8 (1.7) | 39.0 (0.8) |
| 3-4.9 | 23.6 (0.6) | 26.1 (0.8) | 22.5 (1.5) | 23.2 (0.7) |
| >=5 | 26.4 (1.0) | 27.0 (1.0) | 26.5 (1.7) | 24.2 (0.8) |
| Insurance: % (SE) |  |  |  |  |
| None | 22.1 (0.7) | 20.6 (0.8) | 16.4 (1.1) | 11.2 (0.5) |
| Medicaid | 5.8 (0.3) | 4.3 (0.3) | 3.9 (0.5) | 5.3 (0.3) |
| Medicare | 3.4 (0.2) | 6.1 (0.3) | 13.4 (0.9) | 17.9 (0.5) |
| Private | 62.9 (0.9) | 64.2 (1.0) | 60.9 (1.6) | 59.4 (0.8) |
| Other government | 5.8 (0.3) | 4.7 (0.4) | 5.4 (0.7) | 6.2 (0.4) |
| BMI: % (SE) |  |  |  |  |
| <25 | 43.1 (0.7) | 28.3 (0.9) | 27.2 (1.4) | 17.4 (0.4) |
| 25-29.9 | 32.5 (0.6) | 34.4 (0.8) | 34.6 (1.4) | 32.1 (0.6) |
| >=30 | 24.5 (0.6) | 37.3 (0.8) | 38.2 (1.7) | 50.5 (0.7) |
| Comorbidities: Yes % (SE) |  |  |  |  |
| Diabetes mellitus | 3.0 (0.2) | 5.2 (0.4) | 7.0 (0.6) | 20.6 (0.4) |
| High cholesterol | 24.4 (0.7) | 30.3 (0.9) | 33.8 (1.6) | 57.2 (0.7) |
| Coronary heart disease | 1.2 (0.1) | 1.5 (0.2) | 1.8 (0.3) | 8.3 (0.4) |
| Stroke | 0.9 (0.1) | 1.3 (0.1) | 2.2 (0.3) | 6.8 (0.3) |
| Congestive heart failure | 0.7 (0.1) | 0.8 (0.1) | 1.6 (0.3) | 5.9 (0.3) |
| Dietary intake: mean (SE) |  |  |  |  |
| Total fat, % kcal | 33.9 (0.1) | 34.2 (0.2) | 33.6 (0.3) | 34.6 (0.1) |
| Saturated fat, % kcal | 11.0 (0.1) | 11.2 (0.1) | 11.1 (0.1) | 11.3 (0.1) |
| Protein, % kcal | 15.8 (0.1) | 15.7 (0.1) | 15.4 (0.1) | 15.8 (0.1) |
| Fiber, g/1000 kcal | 8.2 (0.1) | 7.9 (0.1) | 8.1 (0.1) | 8.3 (0.1) |
| Cholesterol, mg/1000 kcal | 134 (1) | 138 (2) | 141 (3) | 143 (1) |
| Calcium, mg/1000 kcal | 470 (3) | 448 (3) | 450 (7) | 466 (6) |
| Magnesium, mg/1000 kcal | 150 (1) | 144 (1) | 145 (2) | 150 (2) |
| Potassium, mg/1000 kcal | 1311 (7) | 1283 (8) | 1335 (14) | 1357 (7) |
| Sodium, mg/1000 kcal | 1677 (8) | 1653 (9) | 1657 (16) | 1725 (24) |

Note. Values are survey-weighted percentages or means with standard errors unless otherwise indicated. Hypertension-status categories were mutually exclusive. Measured blood pressure categories were assigned only among adults without self-reported hypertension.

### DASH Score Among Adults with Self-Reported Hypertension

DASH score differed by several sociodemographic and clinical characteristics in multivariable-adjusted models; age- and energy-adjusted estimates were directionally consistent and are presented in Table 3. In multivariable-adjusted models, DASH score was significantly associated with survey period, age group, race/ethnicity, education, BMI category, diabetes mellitus, and stroke status. Adjusted DASH score was lower in 2011–2018 than in 2003–2010 (2.85 [95% CI, 2.78–2.92] vs. 2.98 [95% CI, 2.92–3.05]; P=.007) (Figure 1). Race/ethnicity was strongly associated with multivariable-adjusted DASH score (overall P<.001). Adjusted DASH score was lowest among non-Hispanic Black adults (2.56 [95% CI, 2.48–2.63]) and was 2.89 (95% CI, 2.83–2.96) among non-Hispanic White adults. Clinical characteristics were also associated with multivariable-adjusted DASH score. By BMI category, adjusted DASH score was highest among adults with BMI <25 kg/m² (3.15 [95% CI, 3.04–3.26]), and lowest among adults with BMI ≥30 kg/m² (2.77 [95% CI, 2.72– 2.83]; overall P<.001). Adjusted DASH score was lower among adults with diabetes mellitus than among those without diabetes mellitus (2.81 [95% CI, 2.73–2.90] vs. 2.93 [95% CI, 2.88– 2.99]; P=.026) and lower among adults with prior stroke than among those without prior stroke (2.70 [95% CI, 2.56–2.84] vs. 2.93 [95% CI, 2.88–2.97]; P=.003).

**Figure 1.**
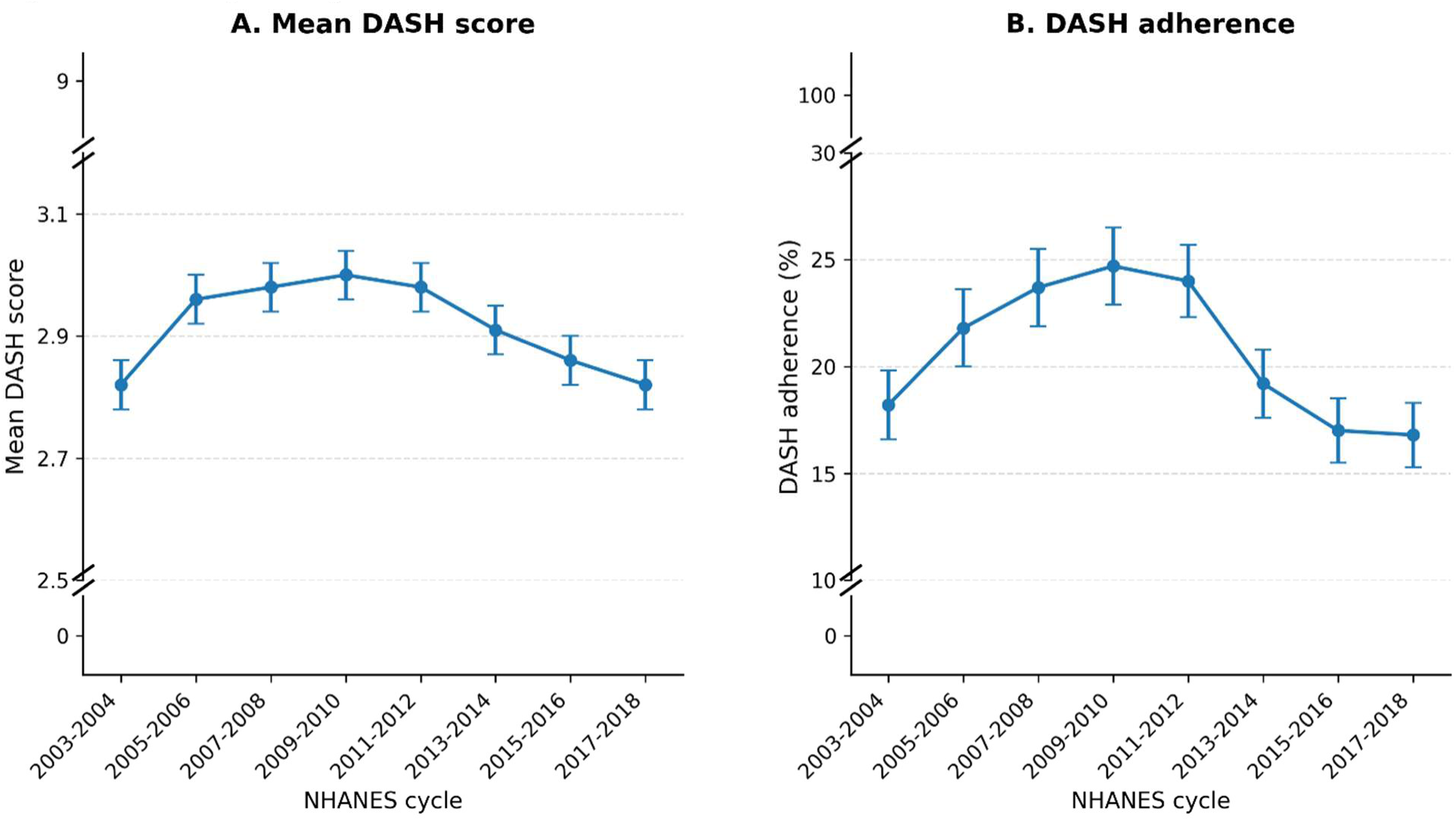
Two-year-cycle DASH score and DASH adherence trends, NHANES 2003–2018. Survey-weighted estimates are shown for U.S. adults with self-reported hypertension across consecutive 2-year NHANES cycles from 2003–2004 through 2017–2018. Panel A shows the observed mean DASH score on a 9-component nutrient-based scale, and Panel B shows the observed percentage of adults classified as DASH accordant, defined as a DASH score ≥4.5. DASH score and DASH adherence increased through 2009–2010 and then declined steadily through 2017–2018. Error bars indicate 95% confidence intervals.

**Table 3.**
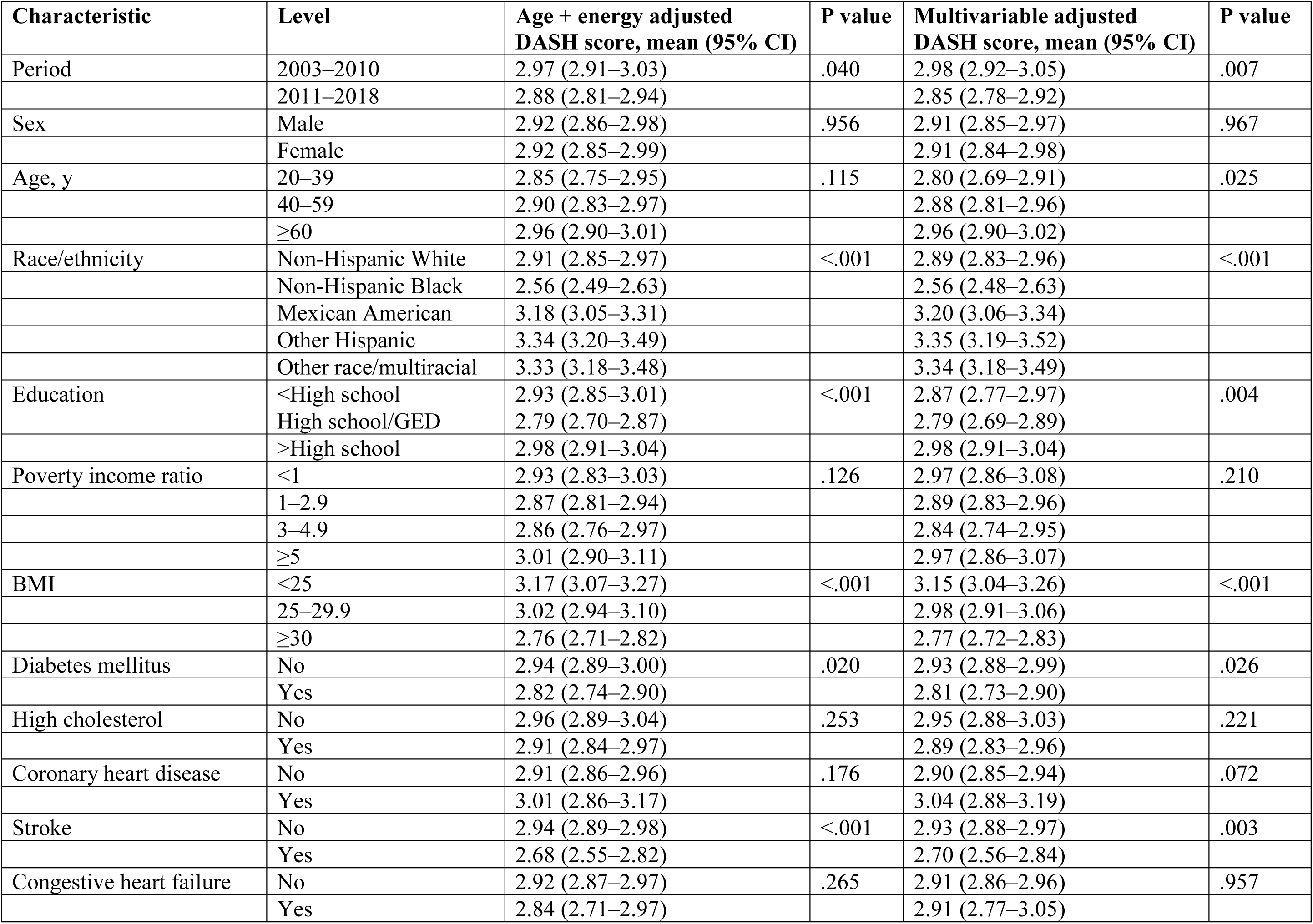
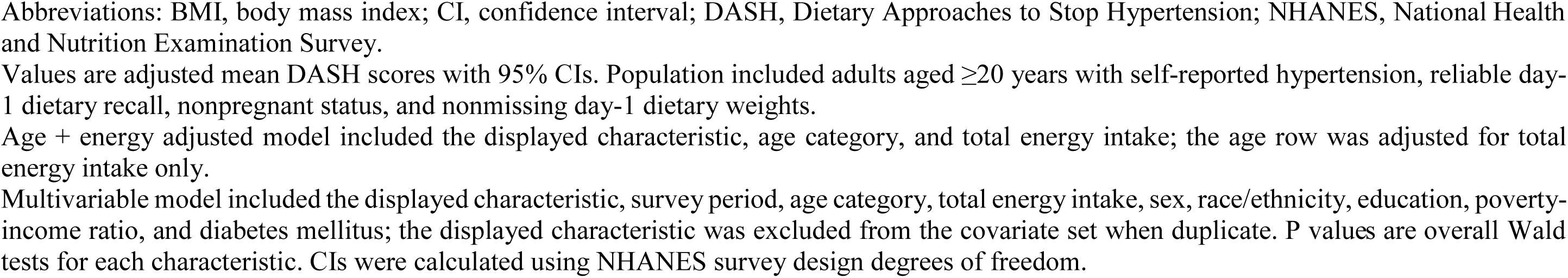
DASH score among adults with self-reported hypertension, NHANES 2003-2018.

| Characteristic | Level | Age + energy adjusted<br>DASH score, mean (95% CI) | P value | Multivariable adjusted<br>DASH score, mean (95% CI) | P value |
| --- | --- | --- | --- | --- | --- |
| Period | 2003–2010 | 2.97 (2.91–3.03) | .040 | 2.98 (2.92–3.05) | .007 |
|  | 2011–2018 | 2.88 (2.81–2.94) |  | 2.85 (2.78–2.92) |  |
| Sex | Male | 2.92 (2.86–2.98) | .956 | 2.91 (2.85–2.97) | .967 |
|  | Female | 2.92 (2.85–2.99) |  | 2.91 (2.84–2.98) |  |
| Age, y | 20–39 | 2.85 (2.75–2.95) | .115 | 2.80 (2.69–2.91) | .025 |
|  | 40–59 | 2.90 (2.83–2.97) |  | 2.88 (2.81–2.96) |  |
|  | ≥60 | 2.96 (2.90–3.01) |  | 2.96 (2.90–3.02) |  |
| Race/ethnicity | Non-Hispanic White | 2.91 (2.85–2.97) | <.001 | 2.89 (2.83–2.96) | <.001 |
|  | Non-Hispanic Black | 2.56 (2.49–2.63) |  | 2.56 (2.48–2.63) |  |
|  | Mexican American | 3.18 (3.05–3.31) |  | 3.20 (3.06–3.34) |  |
|  | Other Hispanic | 3.34 (3.20–3.49) |  | 3.35 (3.19–3.52) |  |
|  | Other race/multiracial | 3.33 (3.18–3.48) |  | 3.34 (3.18–3.49) |  |
| Education | <High school | 2.93 (2.85–3.01) | <.001 | 2.87 (2.77–2.97) | .004 |
|  | High school/GED | 2.79 (2.70–2.87) |  | 2.79 (2.69–2.89) |  |
|  | >High school | 2.98 (2.91–3.04) |  | 2.98 (2.91–3.04) |  |
| Poverty income ratio | <1 | 2.93 (2.83–3.03) | .126 | 2.97 (2.86–3.08) | .210 |
|  | 1–2.9 | 2.87 (2.81–2.94) |  | 2.89 (2.83–2.96) |  |
|  | 3–4.9 | 2.86 (2.76–2.97) |  | 2.84 (2.74–2.95) |  |
|  | ≥5 | 3.01 (2.90–3.11) |  | 2.97 (2.86–3.07) |  |
| BMI | <25 | 3.17 (3.07–3.27) | <.001 | 3.15 (3.04–3.26) | <.001 |
|  | 25–29.9 | 3.02 (2.94–3.10) |  | 2.98 (2.91–3.06) |  |
|  | ≥30 | 2.76 (2.71–2.82) |  | 2.77 (2.72–2.83) |  |
| Diabetes mellitus | No | 2.94 (2.89–3.00) | .020 | 2.93 (2.88–2.99) | .026 |
|  | Yes | 2.82 (2.74–2.90) |  | 2.81 (2.73–2.90) |  |
| High cholesterol | No | 2.96 (2.89–3.04) | .253 | 2.95 (2.88–3.03) | .221 |
|  | Yes | 2.91 (2.84–2.97) |  | 2.89 (2.83–2.96) |  |
| Coronary heart disease | No | 2.91 (2.86–2.96) | .176 | 2.90 (2.85–2.94) | .072 |
|  | Yes | 3.01 (2.86–3.17) |  | 3.04 (2.88–3.19) |  |
| Stroke | No | 2.94 (2.89–2.98) | <.001 | 2.93 (2.88–2.97) | .003 |
|  | Yes | 2.68 (2.55–2.82) |  | 2.70 (2.56–2.84) |  |
| Congestive heart failure | No | 2.92 (2.87–2.97) | .265 | 2.91 (2.86–2.96) | .957 |
|  | Yes | 2.84 (2.71–2.97) |  | 2.91 (2.77–3.05) |  |

Abbreviations: BMI, body mass index; CI, confidence interval; DASH, Dietary Approaches to Stop Hypertension; NHANES, National Health and Nutrition Examination Survey.
Values are adjusted mean DASH scores with 95% CIs. Population included adults aged $\geq 20$ years with self-reported hypertension, reliable day-1 dietary recall, nonpregnant status, and nonmissing day-1 dietary weights.
Age + energy adjusted model included the displayed characteristic, age category, and total energy intake; the age row was adjusted for total energy intake only.
Multivariable model included the displayed characteristic, survey period, age category, total energy intake, sex, race/ethnicity, education, poverty-income ratio, and diabetes mellitus; the displayed characteristic was excluded from the covariate set when duplicate. P values are overall Wald tests for each characteristic. CIs were calculated using NHANES survey design degrees of freedom.

### DASH Score, DASH Adherence, and Nutrient-Target Attainment

In multivariable-adjusted models, mean DASH score was lower in 2011–2018 than in 2003– 2010 (2.85 [SE, 0.04] vs. 2.98 [SE, 0.03]), corresponding to an absolute difference of −0.14 points (P=.007) (Table 4). Age- and energy-adjusted estimates were directionally consistent with multivariable-adjusted findings and are presented in Table 4. Multivariable-adjusted DASH adherence was also lower in 2011–2018 than in 2003–2010 (18.6% [SE, 0.8%] vs. 22.1% [SE, 0.8%]), corresponding to an absolute difference of −3.5 percentage points (P=.003). There was substantial variation in the proportion of adults meeting individual DASH nutrient targets. After multivariable adjustment, statistically significant period differences were observed for total fat and sodium target attainment, whereas differences in saturated fat, protein, cholesterol, fiber, magnesium, calcium, and potassium target attainment were not statistically significant. Target attainment remained low for several DASH components in 2011–2018 (Figure 2).

**Figure 2.**
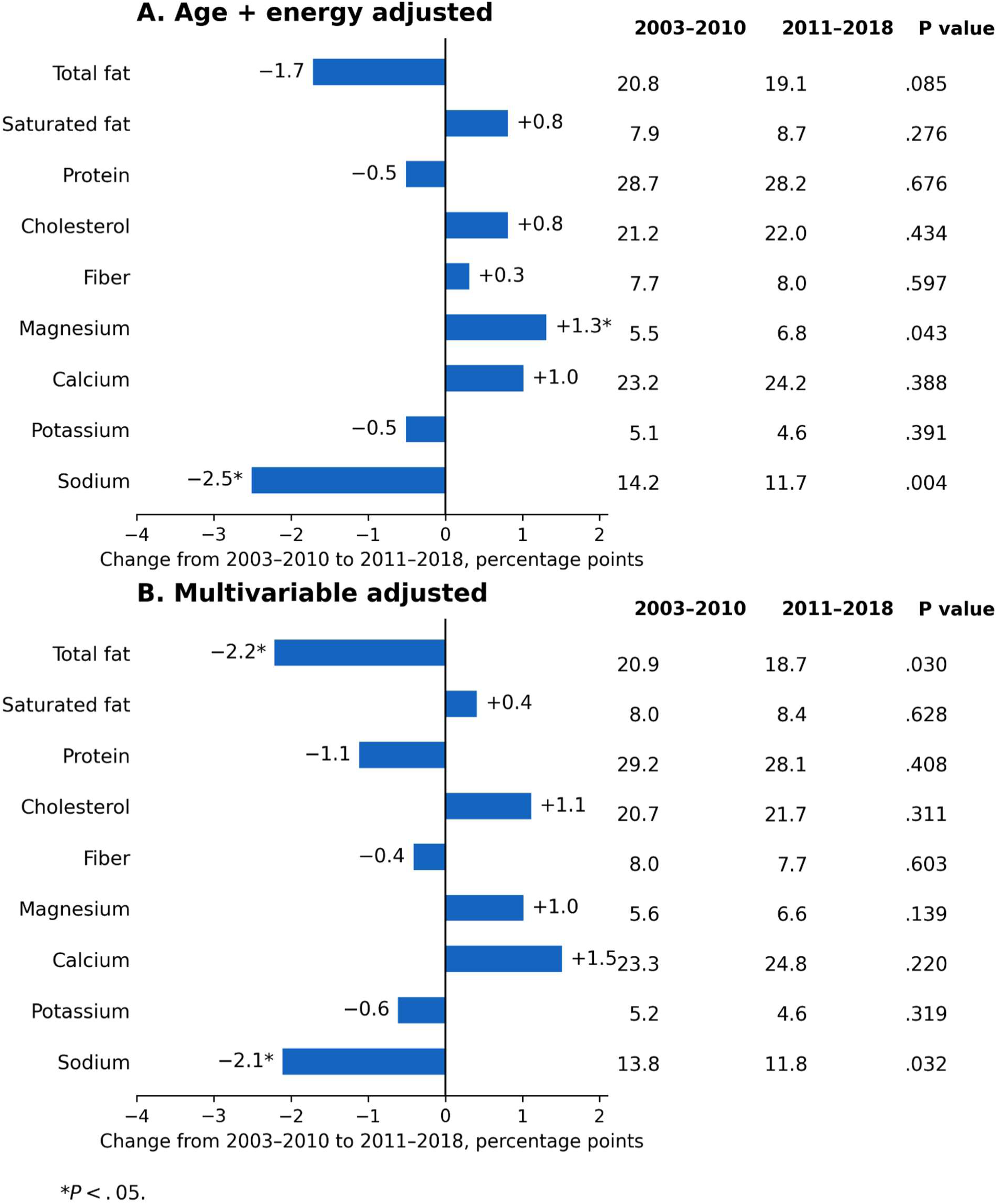
Changes in DASH nutrient-target attainment among U.S. adults with self-reported hypertension, NHANES 2003–2018. Survey-weighted estimates show the percentage of adults meeting each DASH nutrient target in 2003–2010 and 2011–2018. Panel A presents age- and energy-adjusted estimates, and Panel B presents multivariable-adjusted estimates. Bars indicate the percentage-point change from 2003–2010 to 2011–2018; negative values indicate lower target attainment in 2011–2018. P values correspond to the adjusted comparison between survey periods. *P < .05.

**Table 4.** Adjusted mean DASH score, and percent DASH adherence, and nutrient-target achievement among adults with self-reported hypertension, NHANES 2003–2010 vs 2011–2018.

| Outcome | Age + energy adjusted |  |  |  | Multivariable adjusted |  |  |  |
| --- | --- | --- | --- | --- | --- | --- | --- | --- |
|  | 2003–2010 | 2011–2018 | Change | p value | 2003–2010 | 2011–2018 | Change | p value |
| DASH score, mean | 2.97 (0.03) | 2.88 (0.03) | -0.10 | .040 | 2.98 (0.03) | 2.85 (0.04) | -0.14 | .007 |
| DASH adherence, % | 21.8 (0.8) | 19.5 (0.7) | -2.3 | .031 | 22.1 (0.8) | 18.6 (0.8) | -3.5 | .003 |
| Total fat target achieved, % | 20.8 (0.8) | 19.1 (0.6) | -1.7 | .085 | 20.9 (0.7) | 18.7 (0.7) | -2.2 | .030 |
| Saturated fat target achieved, % | 7.9 (0.5) | 8.7 (0.5) | 0.80 | .276 | 8.0 (0.5) | 8.4 (0.6) | 0.40 | .628 |
| Protein target achieved, % | 28.7 (0.9) | 28.2 (0.9) | -0.50 | .676 | 29.2 (1.0) | 28.1 (0.9) | -1.1 | .408 |
| Cholesterol target achieved, % | 21.2 (0.7) | 22.0 (0.7) | 0.80 | .434 | 20.7 (0.8) | 21.7 (0.7) | 1.1 | .311 |
| Fiber target achieved, % | 7.7 (0.4) | 8.0 (0.5) | 0.30 | .597 | 8.0 (0.4) | 7.7 (0.5) | -0.40 | .603 |
| Magnesium target achieved, % | 5.5 (0.4) | 6.8 (0.5) | 1.3 | .043 | 5.6 (0.4) | 6.6 (0.6) | 1.0 | .139 |
| Calcium target achieved, % | 23.2 (0.8) | 24.2 (0.8) | 1.0 | .388 | 23.3 (0.9) | 24.8 (0.8) | 1.5 | .220 |
| Potassium target achieved, % | 5.1 (0.4) | 4.6 (0.4) | -0.50 | .391 | 5.2 (0.4) | 4.6 (0.4) | -0.60 | .319 |
| Sodium target achieved, % | 14.2 (0.7) | 11.7 (0.5) | -2.5 | .004 | 13.8 (0.7) | 11.8 (0.6) | -2.1 | .032 |

**Note.** Values are adjusted estimates with standard errors in parentheses. DASH score is reported as an adjusted mean. DASH adherence and nutrient-target achievement outcomes are reported as adjusted percentages. Change is calculated as 2011–2018 minus 2003–2010; changes for DASH adherence and nutrient-target achievement are percentage-point differences. A+E indicates age- and energy-adjusted; MV, multivariable-adjusted

#### Multivariable Odds of DASH Adherence

In multivariable logistic regression, adults with self-reported hypertension in 2011–2018 had lower odds of DASH adherence than those in 2003– 2010 (OR, 0.78; 95% CI, 0.67–0.91; P=.002) (Table 5). Older age was associated with higher odds of DASH adherence. Compared with adults aged 20–39 years, adults aged 40–59 years had higher odds of DASH adherence (OR, 1.34; 95% CI, 1.02–1.75; P=.034). Race/ethnicity was associated with DASH adherence. Compared with non-Hispanic White adults, non-Hispanic Black adults had lower odds of DASH adherence (OR, 0.70; 95% CI, 0.59–0.84; P<.001).

**Table 5.**
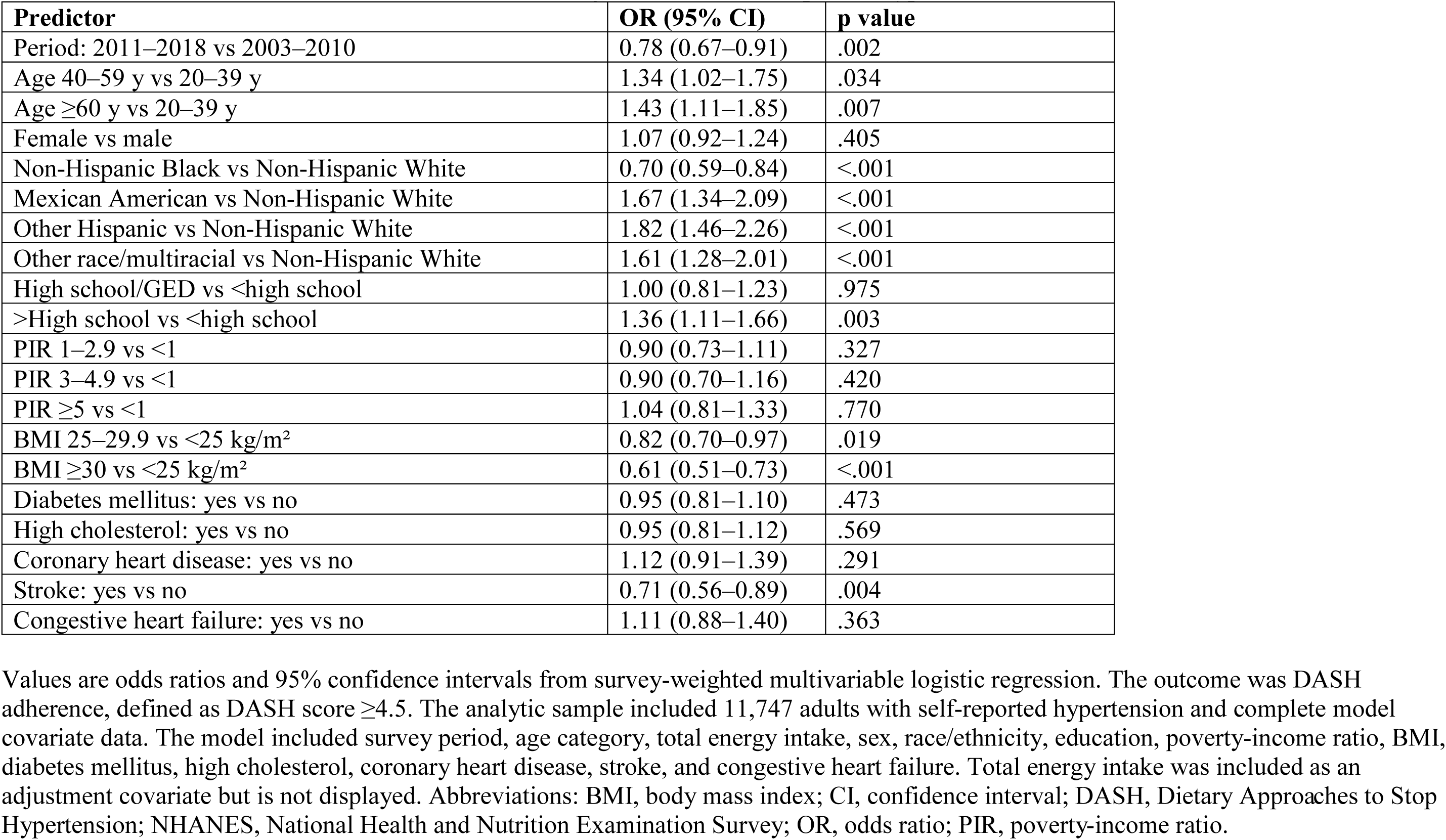
Multivariable odds of DASH adherence among adults with self-reported hypertension, NHANES 2003-2018.

Mexican American adults (OR, 1.67; 95% CI, 1.34–2.09; P<.001), and other Hispanic adults (OR, 1.82; 95% CI, 1.46–2.26; P<.001), had higher odds of DASH adherence. BMI was inversely associated with DASH adherence. Compared with adults with BMI <25 kg/m², adults with BMI 25–29.9 kg/m² had lower odds of DASH adherence (OR, 0.82; 95% CI, 0.70–0.97; P=.02), and adults with BMI ≥30 kg/m² had substantially lower odds (OR, 0.61; 95% CI, 0.51– 0.73; P<.001). Among clinical conditions, stroke was associated with lower odds of DASH adherence (OR, 0.71; 95% CI, 0.56–0.89; P=.004). Diabetes mellitus (OR, 0.95; 95% CI, 0.81– 1.10; P=.47), high cholesterol (OR, 0.95; 95% CI, 0.81–1.12; P=.57), coronary heart disease (OR, 1.12; 95% CI, 0.91–1.39; P=.29), and congestive heart failure (OR, 1.11; 95% CI, 0.88– 1.40; P=.36) were not significantly associated with DASH adherence after multivariable adjustment (Figure 3).

**Figure 3.**
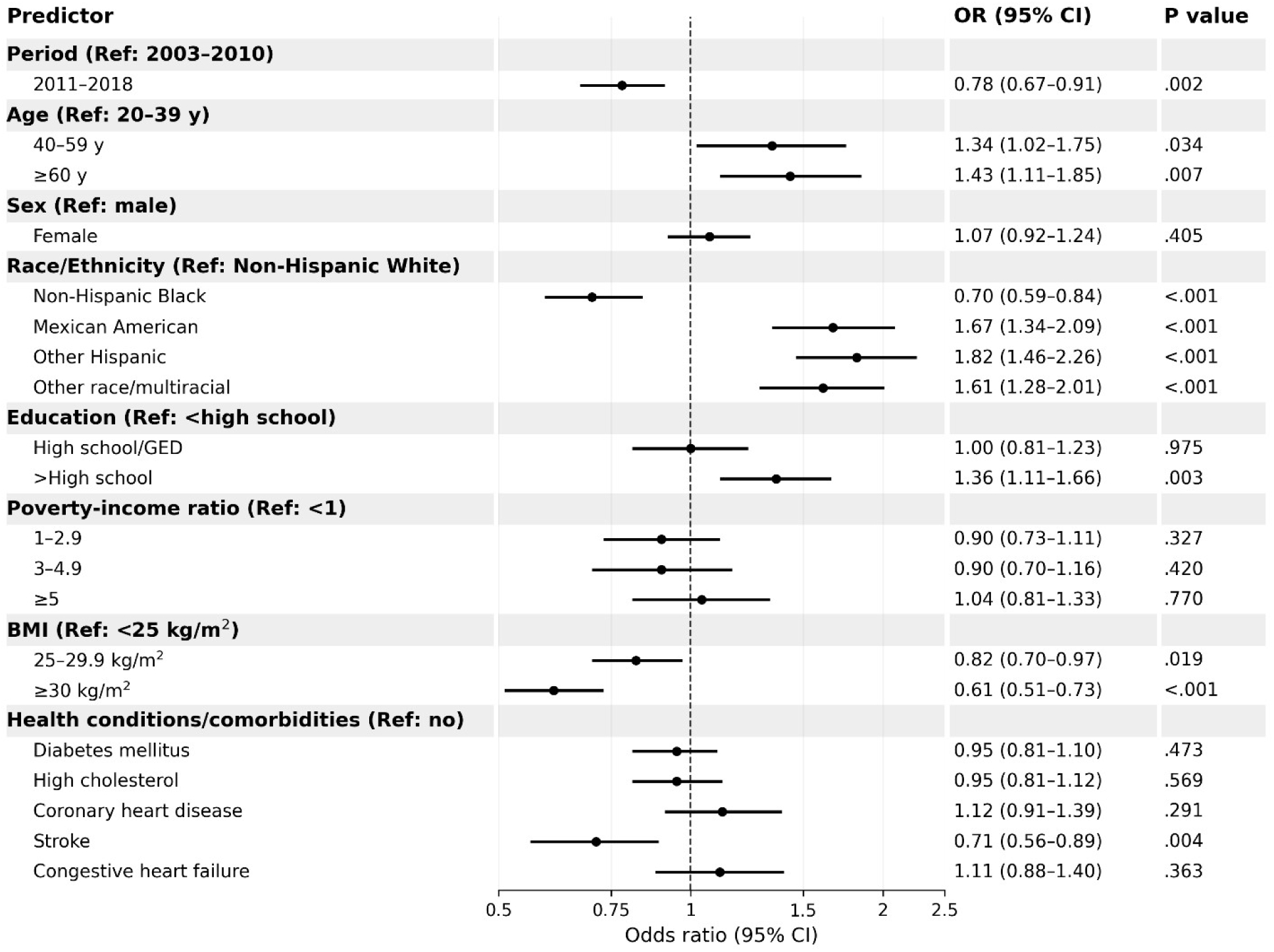
Multivariable predictors of DASH dietary adherence among U.S. adults with self-reported hypertension, NHANES 2003–2018. Adjusted odds ratios and 95% confidence intervals are shown for DASH dietary adherence, defined as a DASH score ≥4.5 on a 9-component nutrient-based scale. Estimates are from a survey-weighted multivariable logistic regression model adjusted for survey period, age category, total energy intake, sex, race/ethnicity, education, poverty-income ratio, body mass index, diabetes mellitus, high cholesterol, coronary heart disease, stroke, and congestive heart failure. The dashed vertical line indicates an odds ratio of 1.0. Values to the left of the line indicate lower odds of DASH adherence, and values to the right indicate higher odds. P values correspond to the adjusted association for each predictor level compared with the reference category.

#### Sensitivity Analysis

Absolute DASH scores and DASH adherence were lower than reported by Mellen et. al, but period differences were directionally consistent with their findings (Table 6). In multivariable-adjusted models, mean DASH score was lower in 2011–2018 than in 2003–2010 (2.34 [SE, 0.03] vs. 2.44 [SE, 0.03]), corresponding to an absolute difference of −0.10 points (p=.025). Multivariable-adjusted DASH adherence was also lower in 2011–2018 than in 2003– 2010 (11.8% [SE, 0.6%] vs. 14.4% [SE, 0.7%]), corresponding to an absolute difference of −2.6 percentage points (p=.007). Age- and energy-adjusted estimates were directionally consistent but did not reach statistical significance and are presented in Table 6.

**Table 6.** Sensitivity analysis using the published 0.5 intermediate-score rule among adults with self-reported hypertension, NHANES 2003–2018.

| Estimate | DASH score |  | DASH adherent |  |
| --- | --- | --- | --- | --- |
|  | Age + Energy adjusted | Multivariate adjusted | Age + Energy adjusted | Multivariate adjusted |
| **2003–2010** | 2.43 (0.03) | 2.44 (0.03) | 13.9 (0.7) | 14.4 (0.7) |
| **2011–2018** | 2.36 (0.03) | 2.34 (0.03) | 12.4 (0.6) | 11.8 (0.6) |
| **Change** | −0.07 | −0.10 | −1.6 | −2.6 |
| **P value** | .096 | .025 | .085 | .007 |

**Notes.** Values are adjusted means or percentages with standard errors. DASH adherence was defined as DASH score $\geq 4.5$ . The primary analysis assigned 0.75 points for intermediate targets; this sensitivity analysis assigned 0.5 points for intermediate targets, consistent with the published scoring rule. Change represents 2011–2018 minus 2003–2010. Abbreviations: DASH, Dietary Approaches to Stop Hypertension; NHANES, National Health and Nutrition Examination Survey.

#### Historical Context Across NHANES Periods

Age- and energy-adjusted DASH score was highest in NHANES III 1988–1994 and lower in each subsequent NHANES period evaluated for historical context (Table 7; Figure 1). Mean DASH score was 3.22 (SE, 0.07) in 1988–1994, 2.92 (SE, 0.05) in 1999–2004, 3.03 (SE, 0.03) in 2005–2010, and 2.88 (SE, 0.03) in 2011–2018. DASH adherence showed a similar overall pattern, with the highest estimate in 1988–1994, lower estimates in 1999–2004 and 2011–2018, and partial recovery in 2005–2010. Two-year-cycle analyses from 2003–2018 provided additional context for the recent-period comparison. Observed DASH score increased through 2009–2010 and then declined through 2017–2018 (Figure 4). Parallel age- and energy-adjusted and multivariable-adjusted historical comparisons of DASH adherence and individual nutrient-target achievement are provided in Supplemental Tables S2A and S2B.

**Figure 4.**
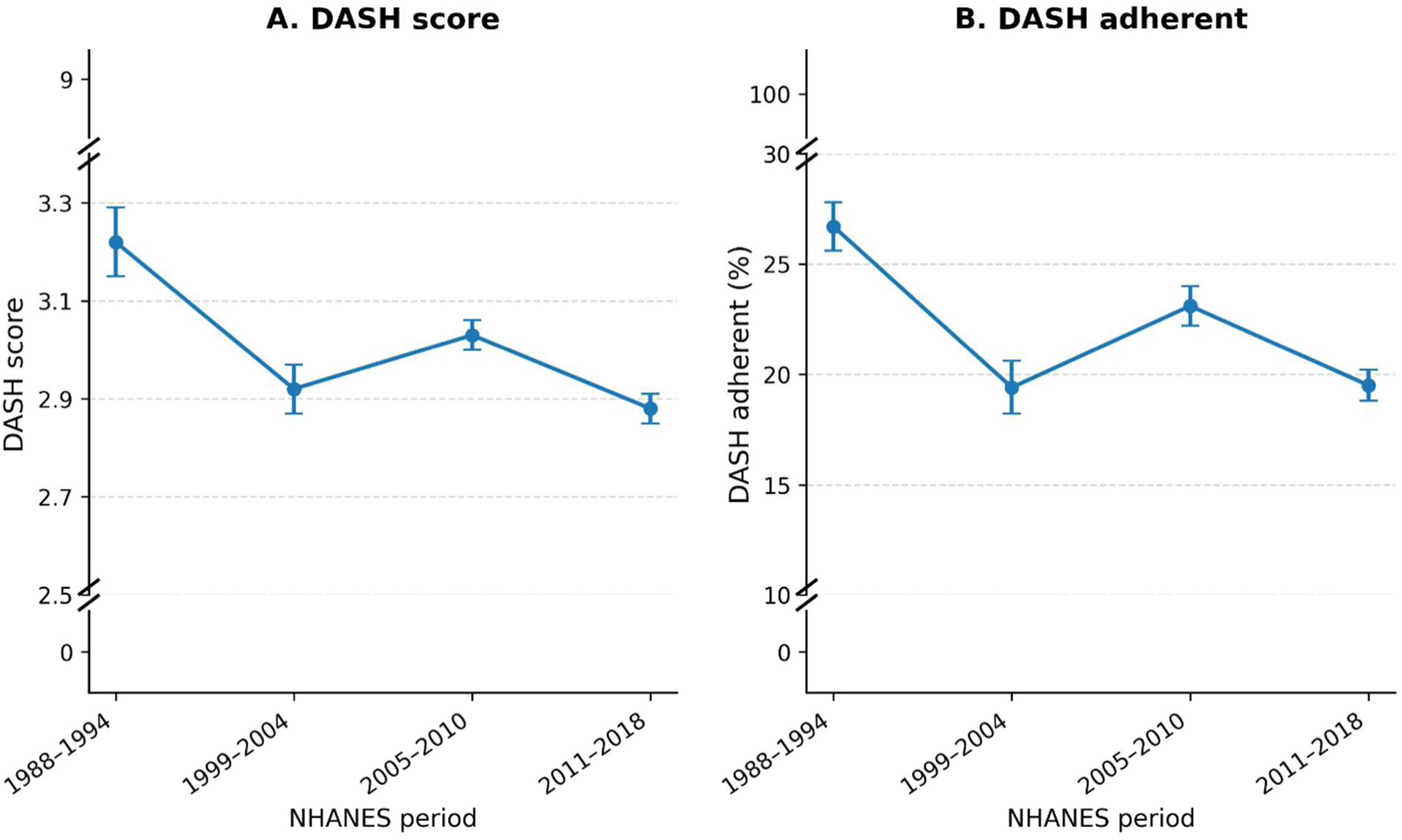
Historical DASH score and DASH adherence trends across four non-overlapping periods. Survey-weighted estimates are shown for U.S. adults with self-reported hypertension across NHANES 1988–1994, 1999–2004, 2005–2010, and 2011–2018. Panel A shows the mean DASH score on a 9-component nutrient-based scale. Panel B shows the percentage of adults meeting the study definition of DASH adherence, defined as a DASH score of at least 4.5. Estimates for 1988–1994 and 1999–2004 are from Mellen et al. [12] and estimates for 2005–2010 and 2011– 2018 were generated in the present extension analysis using comparable non-overlapping NHANES periods. Error bars indicate 95% confidence intervals

**Table 7.**
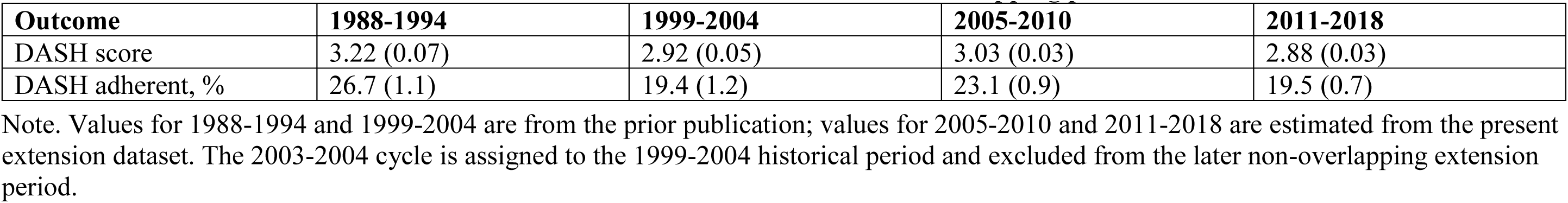
Historical context: DASH score and DASH adherence across four non-overlapping periods.

## DISCUSSION

In this nationally representative study of U.S. adults with self-reported hypertension, DASH dietary adherence remained low from 2003-2010 and worsened between 2011 through 2018. Both mean DASH score and the proportion classified as DASH adherent were lower in 2011– 2018 than in 2003–2010, persisting after adjustment for demographic, socioeconomic, and clinical covariates. These findings extend those of Mellen et al., who documented low and worsening DASH adherence among adults with hypertension from 1988–1994 to 1999–2004 [12], and indicate that this pattern has continued. More than two decades after the original DASH trial and repeated guideline endorsement, the typical diet of U.S. adults with self-reported hypertension remains poorly aligned with the DASH dietary pattern [5,6,10–12].

The observed decline in DASH adherence occurred despite substantial public health attention to dietary patterns and sodium reduction. Successive editions of the Dietary Guidelines for Americans increasingly emphasized overall dietary patterns, with the 2010 and 2015–2020 editions explicitly discussing DASH and the 2020–2025 edition continuing to emphasize nutrient-dense foods and limits on sodium and saturated fat [13–15]. The American Heart Association has similarly identified DASH as highly aligned with its 2021 dietary guidance, while emphasizing that sustained adherence is shaped by clinical, cultural, and environmental factors [16]. National efforts also included voluntary industry sodium targets and menu-labeling requirements for chain restaurants [17,18].

Collectively, these findings suggest that guideline endorsements, voluntary industry initiatives, and information-based labeling policies alone have been insufficient to shift dietary patterns at the population level, reinforcing the need for more comprehensive implementation strategies. These findings are important because self-reported hypertension identifies a large, clinically high-risk population for whom DASH implementation is directly relevant. By 2011–2018, more than 38 million U.S. adults had self-reported hypertension, and compared with normotensive adults, this group had substantially higher prevalence of obesity, diabetes, high cholesterol, and prior cardiovascular conditions. Yet adults with self-reported hypertension had the lowest adjusted DASH scores across hypertension-status categories, suggesting that diagnosis and health care contact alone have not translated into guideline-aligned dietary patterns [1,2,10,11]. Within the self-reported hypertension population, adults with obesity, diabetes, and prior stroke had lower adjusted DASH scores, and adults with obesity and prior stroke had lower odds of DASH adherence suggesting that cardiometabolic risk alone does not translate into improved dietary behavior and that nutrition support needs to be more deliberately integrated into hypertension and chronic disease management [5,6,10,11].

The sociodemographic patterning of DASH adherence further suggests that low adherence is shaped by social and structural factors beyond individual knowledge or motivation. Non-Hispanic Black adults had the lowest multivariable-adjusted DASH score and lower odds of DASH adherence than non-Hispanic White adults, despite bearing a disproportionate burden of hypertension and cardiovascular risk [3,4]. Adults without education beyond high school were also less likely to achieve DASH-concordant diets These differences are consistent with evidence that diet quality and hypertension outcomes are shaped by food access, affordability, neighborhood food environments, food literacy, culturally relevant nutrition support, food marketing, and the composition of the commercial food supply [21–25]. Interventions relying solely on counseling or generic recommendations are unlikely to be sufficient; improving adherence will require practical strategies that make DASH-concordant foods easier to afford, obtain, prepare, and sustain across diverse communities. Grocery stores and pharmacy kiosks represent accessible, community-embedded settings where targeted DASH education could be delivered at the point of food and medication access, offering a scalable approach to reaching populations with persistently low adherence[26].

The nutrient-level findings of our analysis clarify the nature of this adherence gap: the proportions meeting total fat and sodium thresholds declined significantly from 2003–2010 to 2011–2018, while attainment of several other thresholds remained persistently low. In 2011– 2018, fewer than 12% of adults with self-reported hypertension met the fiber, magnesium, potassium, or sodium thresholds, reflecting insufficient intake of fruits, vegetables, legumes, whole grains, nuts, and low-fat dairy and mirroring broader U.S. dietary trends [13–15,20].

Sodium reduction remains important, but narrowly focused interventions may be insufficient without also supporting adoption of the full DASH pattern. DASH-focused digital interventions have shown generally favorable effects on blood pressure, diet quality, sodium intake, and weight, though evidence remains heterogeneous [27,28]. The GoFresh randomized trial demonstrated that home-delivered DASH-patterned groceries with dietitian counseling reduced blood pressure and low-density lipoprotein cholesterol among Black adults with hypertension, though benefits were not sustained after the intervention ended, underscoring the need for approaches that maintain support over time [29]. Artificial Intelligence (AI)-enabled tools including large language model-based dietary coaching, automated meal planning, and adaptive messaging may help deliver personalized DASH guidance where dietitian access is limited.

However, current large language model applications primarily inform rather than reliably guide care delivery [30], and nutrition-specific evaluations underscore the need to assess reliability, readability, safety, and clinical oversight before deployment [31]. Future DASH implementation efforts should combine culturally adaptable food substitutions, practical meal-planning support, integration of nutrition services into hypertension care, and linkage to resources reducing cost and access barriers for marginalized subgroups. Additionally, using longitudinal designs, multiple dietary recalls, measured blood pressure, and food-group-based DASH scoring could provide more precise estimates of individual dietary change and its clinical associations.

Research evaluating DASH food planning interventions delivered through community-accessible settings are urgently needed to support sustained and scalable dietary adherence across diverse populations.

## STRENGTHS

This study has several strengths. It extends the original Mellen et al. analysis using nationally representative NHANES data through 2018, applies a comparable DASH scoring framework based on a well-defined and extensively studied dietary pattern, and provides updated population-level evidence on DASH adherence among U.S. adults with self-reported hypertension. The analysis also places contemporary DASH adherence in a longer-term historical context, examines adherence across demographic, socioeconomic, adiposity, and clinical subgroups, and evaluates individual nutrient targets to identify which components of the DASH pattern remain least commonly achieved.

## LIMITATIONS

Several limitations should be considered. NHANES is cross-sectional; therefore, observed differences across survey periods reflect population-level trends rather than individual dietary change. Dietary intake was assessed using a single 24-hour recall, which supports group-level estimates of mean intake but may misclassify individual usual intake and DASH adherence, with potential bias in either direction. Self-reported hypertension may not capture all affected adults, particularly those undiagnosed. Race/ethnicity was categorized using harmonized variables available across the analytic period, limiting assessment of groups not consistently identified across cycles, including non-Hispanic Asian adults.

## CONCLUSIONS

In this nationally representative study of U.S. adults with self-reported hypertension, DASH dietary adherence remained low and declined from 2003–2010 to 2011–2018. Both mean DASH score and the proportion classified as DASH adherent were lower in 2011–2018 than in 2003– 2010, indicating that expanded evidence, guidelines, and public-facing DASH resources have not translated into improved population-level adherence. These findings underscore the need for more effective implementation strategies that address the full DASH dietary pattern, including scalable, culturally adaptable approaches that may include validated digital and AI-assisted dietary planning tools.

## Data Availability

The data analyzed in this study are publicly available from the National Health and Nutrition Examination Survey (NHANES), conducted by the National Center for Health Statistics, including questionnaire, examination, laboratory, and dietary data files and documentation for the relevant survey cycles. NHANES public-use datasets can be accessed through the CDC/NCHS NHANES website

## Author Contributions

VP: Conceptualization, methodology, data curation, writing: original draft, review & editing; AP: Writing: review & editing; DM and CG: Conceptualization, visualization, results interpretation, writing: review & editing.

## SUPPLEMENT

**Supplemental Table S1.** Adjusted DASH score by hypertension status, NHANES 2003-2018.

| Adjustment | Hypertension status | DASH score | Overall P value |
| --- | --- | --- | --- |
| Age + energy | Normotensive | 3.05 (0.03) | <.001 |
|  | Prehypertension | 2.92 (0.03) |  |
|  | Undiagnosed hypertension | 2.88 (0.05) |  |
|  | Self-reported hypertension | 2.81 (0.02) |  |
| Multivariable | Normotensive | 3.01 (0.03) | <.001 |
|  | Prehypertension | 2.90 (0.03) |  |
|  | Undiagnosed hypertension | 2.87 (0.05) |  |
|  | Self-reported hypertension | 2.84 (0.03) |  |

Note. Values are adjusted DASH score means with standard errors. P values are for the overall difference across hypertension-status categories.

**Supplemental Table S2A.** Age- and energy-adjusted DASH adherence and individual nutrient-target achievement among adults with self-reported hypertension across historical NHANES periods.

| **Outcome** | **1988–1994** | **1999–2004** | **2005–2010** | **2011–2018** |
| --- | --- | --- | --- | --- |
| DASH adherence | 26.7 (1.1) | 19.4 (1.2) | 23.2 (0.9) | 19.5 (0.7) |
| Total fat | 39.3 (1.8) | 31.2 (1.7) | 21.8 (0.8) | 19.1 (0.6) |
| Saturated fat | 18.2 (1.2) | 16.5 (1.0) | 7.9 (0.5) | 8.6 (0.5) |
| Protein | 35.3 (1.7) | 38.7 (1.6) | 29.7 (1.1) | 28.1 (0.9) |
| Cholesterol | 30.5 (1.9) | 27.6 (1.5) | 20.7 (0.8) | 22.0 (0.7) |
| Fiber | 16.4 (0.8) | 10.7 (0.7) | 8.1 (0.5) | 8.0 (0.5) |
| Magnesium | 13.6 (1.2) | 6.3 (0.6) | 6.1 (0.4) | 6.8 (0.5) |
| Calcium | 19.3 (1.3) | 21.1 (1.6) | 25.0 (1.0) | 24.2 (0.8) |
| Potassium | 13.3 (1.0) | 11.4 (0.9) | 5.2 (0.5) | 4.6 (0.4) |
| Sodium | 18.6 (1.5) | 16.1 (1.3) | 13.2 (0.7) | 11.7 (0.5) |

*Note.* Values are age- and energy-adjusted percentages (SE). Values for 1988–1994 and 1999–2004 are from the prior publication. Values for 2005–2010 and 2011–2018 were estimated from the present extension dataset using non-overlapping historical periods. For this historical-context analysis, the 2003–2004 cycle is included in the 1999–2004 period, consistent with the prior publication; the 2005–2010 period begins with the 2005–2006 cycle. Abbreviations: DASH, Dietary Approaches to Stop Hypertension; NHANES, National Health and Nutrition Examination Survey; SE, standard error.

**Supplemental Table S2B.** Multivariable-adjusted DASH adherence and individual nutrient-target achievement among adults with self-reported hypertension across historical NHANES periods.

| **Outcome** | **1988–1994** | **1999–2004** | **2005–2010** | **2011–2018** |
| --- | --- | --- | --- | --- |
| DASH adherence | 29.3 (1.5) | 21.7 (1.3) | 23.4 (1.0) | 18.7 (0.8) |
| Total fat | 42.9 (1.8) | 35.9 (2.0) | 21.7 (0.8) | 18.7 (0.7) |
| Saturated fat | 20.6 (1.2) | 20.4 (1.4) | 7.9 (0.5) | 8.5 (0.6) |
| Protein | 43.7 (2.0) | 47.7 (1.9) | 30.2 (1.1) | 28.1 (0.9) |
| Cholesterol | 26.4 (2.2) | 24.3 (1.6) | 20.0 (0.9) | 21.8 (0.7) |
| Fiber | 20.2 (1.5) | 12.3 (0.9) | 8.5 (0.5) | 7.7 (0.5) |
| Magnesium | 14.2 (1.3) | 6.4 (0.8) | 6.2 (0.5) | 6.7 (0.6) |
| Calcium | 19.0 (1.6) | 17.6 (2.0) | 25.1 (1.1) | 24.8 (0.8) |
| Potassium | 12.7 (0.9) | 11.7 (0.9) | 5.3 (0.5) | 4.6 (0.4) |
| Sodium | 17.8 (1.5) | 14.6 (1.3) | 12.6 (0.8) | 11.8 (0.6) |

*Note.* Values are multivariable-adjusted percentages (SE). Multivariable models adjusted for age category, total energy intake, sex, race/ethnicity, education, poverty-income ratio, and diabetes mellitus. Values for 1988–1994 and 1999–2004 are from the prior publication. Values for 2005–2010 and 2011–2018 were estimated from the present extension dataset using non-overlapping historical periods. For this historical-context analysis, the 2003–2004 cycle is included in the 1999–2004 period, consistent with the prior publication; the 2005–2010 period begins with the 2005–2006 cycle. Abbreviations: DASH, Dietary Approaches to Stop Hypertension; NHANES, National Health and Nutrition Examination Survey; SE, standard error.

**Supplemental Table S3.**
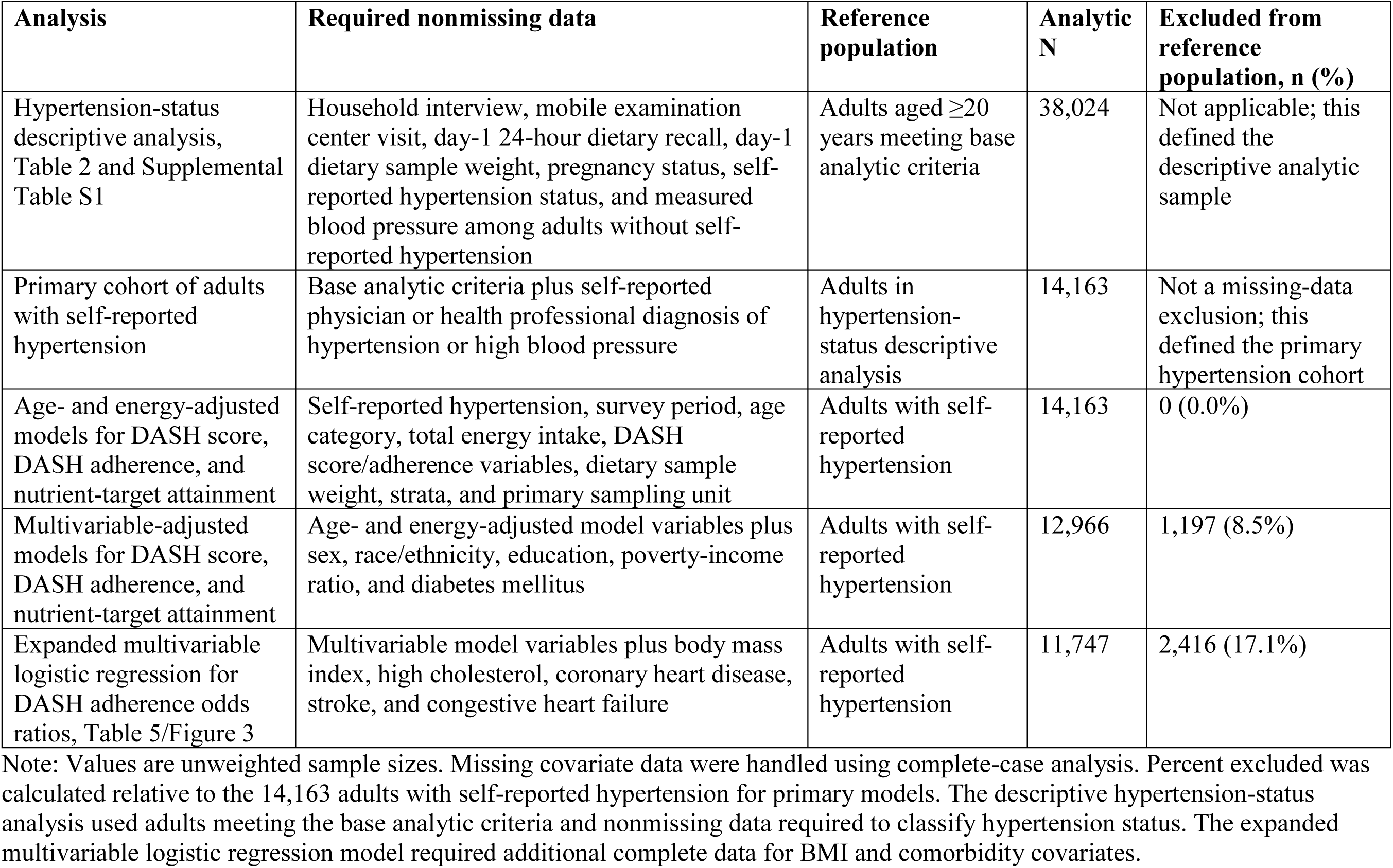
Analytic sample size and missing covariate handling by analysis.

| **Analysis** | **Required nonmissing data** | **Reference population** | **Analytic N** | **Excluded from reference population, n (%)** |
| --- | --- | --- | --- | --- |
| Hypertension-status descriptive analysis, Table 2 and Supplemental Table S1 | Household interview, mobile examination center visit, day-1 24-hour dietary recall, day-1 dietary sample weight, pregnancy status, self-reported hypertension status, and measured blood pressure among adults without self-reported hypertension | Adults aged $\geq 20$ years meeting base analytic criteria | 38,024 | Not applicable; this defined the descriptive analytic sample |
| Primary cohort of adults with self-reported hypertension | Base analytic criteria plus self-reported physician or health professional diagnosis of hypertension or high blood pressure | Adults in hypertension-status descriptive analysis | 14,163 | Not a missing-data exclusion; this defined the primary hypertension cohort |
| Age- and energy-adjusted models for DASH score, DASH adherence, and nutrient-target attainment | Self-reported hypertension, survey period, age category, total energy intake, DASH score/adherence variables, dietary sample weight, strata, and primary sampling unit | Adults with self-reported hypertension | 14,163 | 0 (0.0%) |
| Multivariable-adjusted models for DASH score, DASH adherence, and nutrient-target attainment | Age- and energy-adjusted model variables plus sex, race/ethnicity, education, poverty-income ratio, and diabetes mellitus | Adults with self-reported hypertension | 12,966 | 1,197 (8.5%) |
| Expanded multivariable logistic regression for DASH adherence odds ratios, Table 5/Figure 3 | Multivariable model variables plus body mass index, high cholesterol, coronary heart disease, stroke, and congestive heart failure | Adults with self-reported hypertension | 11,747 | 2,416 (17.1%) |

Note: Values are unweighted sample sizes. Missing covariate data were handled using complete-case analysis. Percent excluded was calculated relative to the 14,163 adults with self-reported hypertension for primary models. The descriptive hypertension-status analysis used adults meeting the base analytic criteria and nonmissing data required to classify hypertension status. The expanded multivariable logistic regression model required additional complete data for BMI and comorbidity covariates.

## References

1. Centers for Disease Control and Prevention. Heart Disease Facts. Atlanta, GA: Centers for Disease Control and Prevention; 2024. Accessed July 5, 2026. https://www.cdc.gov/heart-disease/data-research/facts-stats/index.html

2. Centers for Disease Control and Prevention. High Blood Pressure Facts. Atlanta, GA: Centers for Disease Control and Prevention; 2026. Accessed July 5, 2026. https://www.cdc.gov/high-blood-pressure/data-research/facts-stats/index.html

3. Aggarwal R, Chiu N, Wadhera RK, Moran AE, Raber I, Shen C, Yeh RW, Kazi DS. Racial/ethnic disparities in hypertension prevalence, awareness, treatment, and control in the United States, 2013 to 2018. Hypertension. 2021;78(6):1719–1726. doi:10.1161/HYPERTENSIONAHA.121.17570

4. Chaturvedi A, Zhu A, Gadela NV, Prabhakaran D, Jafar TH. Social determinants of health and disparities in hypertension and cardiovascular diseases. Hypertension. 2024;81(3):387–399. doi:10.1161/HYPERTENSIONAHA.123.21354

5. Appel LJ, Moore TJ, Obarzanek E, Vollmer WM, Svetkey LP, Sacks FM, et al. A clinical trial of the effects of dietary patterns on blood pressure. N Engl J Med. 1997;336(16):1117–1124. doi:10.1056/NEJM199704173361601

6. Sacks FM, Svetkey LP, Vollmer WM, Appel LJ, Bray GA, Harsha D, et al. Effects on blood pressure of reduced dietary sodium and the Dietary Approaches to Stop Hypertension (DASH) diet. N Engl J Med. 2001;344(1):3–10. doi:10.1056/NEJM200101043440101

7. Saneei P, Salehi-Abargouei A, Esmaillzadeh A, Azadbakht L. Influence of Dietary Approaches to Stop Hypertension (DASH) diet on blood pressure: a systematic review and meta-analysis on randomized controlled trials. Nutr Metab Cardiovasc Dis. 2014;24(12):1253–1261. doi:10.1016/j.numecd.2014.06.008

8. Siervo M, Lara J, Chowdhury S, Ashor A, Oggioni C, Mathers JC. Effects of the Dietary Approach to Stop Hypertension (DASH) diet on cardiovascular risk factors: a systematic review and meta-analysis. Br J Nutr. 2015;113(1):1–15. doi:10.1017/S0007114514003341

9. Fung TT, Chiuve SE, McCullough ML, Rexrode KM, Logroscino G, Hu FB. Adherence to a DASH-style diet and risk of coronary heart disease and stroke in women. Arch Intern Med. 2008;168(7):713–720. doi:10.1001/archinte.168.7.713

10. Whelton PK, Carey RM, Aronow WS, Casey DE Jr, Collins KJ, Dennison Himmelfarb C, et al. 2017 ACC/AHA/AAPA/ABC/ACPM/AGS/APhA/ASH/ASPC/NMA/PCNA guideline for the prevention, detection, evaluation, and management of high blood pressure in adults. Hypertension. 2018;71(6):e13–e115. doi:10.1161/HYP.0000000000000065

11. Arnett DK, Blumenthal RS, Albert MA, Buroker AB, Goldberger ZD, Hahn EJ, et al. 2019 ACC/AHA guideline on the primary prevention of cardiovascular disease: a report of the American College of Cardiology/American Heart Association Task Force on Clinical Practice Guidelines. Circulation. 2019;140(11):e596–e646. doi:10.1161/CIR.0000000000000678

12. Mellen PB, Gao SK, Vitolins MZ, Goff DC Jr. Deteriorating dietary habits among adults with hypertension: DASH dietary adherence, NHANES 1988-1994 and 1999-2004. Arch Intern Med. 2008;168(3):308–314. doi:10.1001/archinternmed.2007.119

13. U.S. Department of Agriculture; U.S. Department of Health and Human Services. Dietary Guidelines for Americans, 2010. 7th ed. Washington, DC: U.S. Government Printing Office; 2010.

14. U.S. Department of Health and Human Services; U.S. Department of Agriculture. 2015-2020 Dietary Guidelines for Americans. 8th ed. Washington, DC: U.S. Department of Health and Human Services and U.S. Department of Agriculture; 2015.

15. U.S. Department of Agriculture; U.S. Department of Health and Human Services. Dietary Guidelines for Americans, 2020-2025. 9th ed. Washington, DC: U.S. Department of Agriculture and U.S. Department of Health and Human Services; 2020.

16. Gardner CD, Vadiveloo MK, Petersen KS, Anderson CAM, Springfield S, Van Horn L, Khera A, Lamendola C, Mayo SM, Joseph JJ; American Heart Association Council on Lifestyle and Cardiometabolic Health. Popular dietary patterns: alignment with American Heart Association 2021 dietary guidance: a scientific statement from the American Heart Association. Circulation. 2023;147(22):1715–1730. doi:10.1161/CIR.0000000000001146

17. U.S. Food and Drug Administration. Voluntary Sodium Reduction Goals: Target Mean and Upper Bound Concentrations for Sodium in Commercially Processed, Packaged, and Prepared Foods: Guidance for Industry. Silver Spring, MD: U.S. Food and Drug Administration; 2021.

18. Food and Drug Administration, HHS. Food labeling; nutrition labeling of standard menu items in restaurants and similar retail food establishments. Final rule. Fed Regist. 2014;79(230):71155–71259.

19. Grummon AH, Petimar J, Soto MJ, Bleich SN, Simon D, Cleveland LP, Musicus AA, Novak NL, Block JP. Changes in calorie content of menu items at large chain restaurants after implementation of calorie labels. JAMA Netw Open. 2021;4(12):e2141353. doi:10.1001/jamanetworkopen.2021.41353

20. Juul F, Parekh N, Martinez-Steele E, Monteiro CA, Chang VW. Ultra-processed food consumption among US adults from 2001 to 2018. Am J Clin Nutr. 2022;115(1):211–221. doi:10.1093/ajcn/nqab305

21. Whitt-Glover MC, Hunter JC, Foy CG, Quandt SA, Vitolins MZ, Leng I, Hornbuckle LM, Sanya KA, Bertoni AG. Translating the Dietary Approaches to Stop Hypertension (DASH) diet for use in underresourced, urban African American communities, 2010. Prev Chronic Dis. 2013;10:120088. doi:10.5888/pcd10.120088

22. Bertoni AG, Foy CG, Hunter JC, Quandt SA, Vitolins MZ, Whitt-Glover MC. A multilevel assessment of barriers to adoption of Dietary Approaches to Stop Hypertension (DASH) among African Americans of low socioeconomic status. J Health Care Poor Underserved. 2011;22(4):1205–1220. doi:10.1353/hpu.2011.0142

23. White BM, OoNorasak K, Sesay NA, Haskins D, Robinson CM. Economic and cultural assessment of the DASH eating plan for low-income African Americans: an integrative review. Int J Environ Res Public Health. 2024;21(11):1480. doi:10.3390/ijerph21111480

24. Chin, L., McCormick, J. B., & Greenberg, R. S. (2018). Convening a digitally enabled ecosystem to address the chronic disease burden of an underserved community. NEJM Catalyst, 4(5)

25. Boyland E, Muc M, Coates A, Ells L, Halford JCG, Hill Z, et al. Food marketing, eating and health outcomes in children and adults: a systematic review and meta-analysis. Br J Nutr. 2025;133(6):781–805. doi:10.1017/S0007114524000102.

26. Musicus AA, Kraak VI, Bleich SN. Policy progress in reducing sodium in the American diet, 2010–2019. Annu Rev Nutr. 2020;40:407–435. doi:10.1146/annurev-nutr-122319-040249.

27. Katz ME, Ogungbe O, Commodore-Mensah Y, Berra K, Joseph JJ, Ogedegbe G, et al. Digital health interventions for hypertension management in US populations experiencing health disparities: a systematic review and meta-analysis. JAMA Netw Open. 2024;7(2):e2356070. doi:10.1001/jamanetworkopen.2023.56070

28. Dodge E, Kelly PJ, Aboul-Enein BH. Digital-based nutrition interventions employing the Dietary Approaches to Stop Hypertension (DASH) diet: a systematic scoping review. Int J Hypertens. 2025;2025:6175223. doi:10.1155/ijhy/6175223

29. Juraschek SP, Col H, Ferro K, Turkson-Ocran RAN, Cluett JL, Davis RB, et al. DASH-patterned groceries and effects on blood pressure: the GoFresh randomized clinical trial. JAMA. 2026;335(1):36–48. doi:10.1001/jama.2025.21112

30. Yim D, Khuntia J, Parameswaran V, Meyers A. Preliminary evidence of the use of generative AI in health care clinical services: systematic narrative review. JMIR Med Inform. 2024;12:e52073. doi:10.2196/52073

31. Parameswaran V, Bernard J, Bernard A, Deo N, Tsung S, Lyytinen K, Sharp C, Rodriguez F, Maron DJ, Dash R. Evaluating large language models and retrieval-augmented generation enhancement for delivering guideline-adherent nutrition information for cardiovascular disease prevention: cross-sectional study. J Med Internet Res. 2025;27:e78625. doi:10.2196/78625

